# Feasibility, Usability, and Acceptability of a Novel Open-source Manual Standing Wheelchair in Low Resource Settings

**DOI:** 10.1101/2025.03.05.25323410

**Authors:** Farjana Taoheed, Shovan Parvez, Md Akhlasur Rahman, Muzaffor Hossain, Mohammad Anwar Hossain, Md. Obaidul Haque, Valerie Ann Taylor, Mohammad Sohrab Hossain, Monzurul Alam

**Affiliations:** Centre for the Rehabilitation of the Paralysed, Savar, Bangladesh; Bangladesh Health Professions Institute, Savar, Bangladesh; The University of Sydney, New South Wales, Australia

**Keywords:** Paralysis, standing wheelchair, feasibility, usability, acceptability, low-cost, open

## Abstract

**Background:** Standing wheelchairs significantly improve mobility and independence, but their high-cost limits accessibility. To address this, we developed an low-cost manual standing wheelchair aimed at enhancing independence and quality of life for individuals with paralysis. This study evaluates the feasibility, usability, and acceptability of the wheelchair among individuals in low-economic settings.

**Methods:** Thirty paraplegic wheelchair users from the Centre for the Rehabilitation of the Paralysed, Savar, Bangladesh were purposively selected to assess the wheelchair’s usability. A wheelchair usability questionnaire using a Likert scale was administered, and qualitative feedback was gathered through an acceptability questionnaire.

**Results:** The material cost of the locally developed standing wheelchair was US$166, significantly lower than the median income in Bangladesh, making it affordable for most paraplegics. The study participants (26 males, 4 females; mean age: 30.8 ± 8.5 years) had a mean height of 165.02 ± 6.8 cm and weight of 57.3 ± 8.3 kg. They sustained a complete motor-injury (ASIA A/B) between T4-L2 spinal levels. Participants spent an average of 7.7 ± 1.2 hours per day using the wheelchair. Most participants felt safe (mean score: 8.4/10) and experienced minimal effort in operation (3.3/10). They reported increased confidence (7.7/10) and happiness (8.2/10) when standing, with a lower level of nervousness (2.4/10). Users anticipated increased involvement in household activities (7.3/10), and overall satisfaction was high (7.1/10).

**Conclusions:** The manual standing wheelchair is affordable, easy to operate, and well-accepted by users in rehabilitation settings. Further evaluation in community settings is recommended to confirm its broader feasibility and impact.

## INTRODUCTION

According to a WHO report (2022), out of the 80 million people who need wheelchairs, only 5– 35% globally have access, and only 3% of those are in low-income countries (1). This unmet supply is mostly due to the high cost of wheelchairs (1).

Many neurological conditions result in partial or complete paralysis, requiring the use of a wheelchair for mobility purposes (2). This includes Multiple sclerosis (MS), cerebral palsy (CP), brain injury and individuals with spinal cord injury (SCI) with 29% of individuals with MS, 41% of individuals with CP using wheelchairs indoors and outdoors, and may require a wheelchair for all types of mobility (3).

Over eight hours of sitting a day raises death rates; nevertheless, standing is a healthy substitute that can lower adult mortality (4). Non-ambulatory adults with neurological conditions like stroke, SCI, brain injury or MS often sit for over 8 hours a day, leading to secondary complications including body structure and functions such as altered muscle tone or spasticity, range of motion (ROM) limitations or contractures, muscle weakness, constipation, decreased bone mineral density (BMD) with increased risk for fractures and bone pain, as well as activity limitations and participation restrictions (4).

Several health-related improvements are reported due to standing among persons with neurological conditions such as brain injury, stroke, MS, SCI and others (5, 6). It has been documented that standing provides various health and psychological benefits as well as preventing secondary complications (7). Standing properly is important for a variety of daily living (ADL) tasks as well as for maintaining a range of health advantages (5). Standing helps individuals with SCI with their spasticity, bowel and bladder function, circulation, and life satisfaction (8). Additionally, data suggests that people with SCI who sometimes stand have fewer pressure ulcers. The standing posture relieves pressure from the sacrum, buttocks, and ischial tuberosity (9). Standing may also stop the decrease of bone density in the lower limbs (10). According to research on bone health, practicing standing for 60 minutes a day is the greatest way to increase bone mineral density (11). Standing has also been associated with improved skin integrity, higher muscular strength, and range of motion, improved respiratory function, a decreased risk of contractures, and skeletal anomalies (9). Additionally, a recent study discovered that standing was particularly advantageous for a person with SCI in terms of functional reach, digestion, sleep, pain, fatigue, eye-level social engagement, and simply feeling good. Overall standing posture has a wide range of psychosocial and quality of life (QoL) advantages (9). Posture transformation from sitting to standing independently enhances job performance, employment prospects, and blood circulation, among other benefits (12). Standing five days per week for 60 minutes each day improves bone density and hip stability, while a range of motion in the lower extremities requires 45 minutes, and spasticity reduction can occur within 30 minutes (13).

Standing devices currently exist in three forms: standing frames, manual standing wheelchairs and powered standing wheelchairs (14). Standing frames are versatile tools that assist individuals with various disabilities in achieving or maintaining a standing posture but users dislike standing in a frame that never moves (15). Some standing frames provide limited mobility in a standing position and wheelchair use is not intended for these frames (14). Moreover, individuals with disabilities often struggle with independent standing. On the other hand, Power standing wheelchairs are expensive, difficult to transport, and restrict the chance of incidental exercise that occurs with daily manual wheelchair propulsion (14). Side by side some standing frames are integrated into the manual wheelchair to facilitate the users to avoid transferring to a standing frame (12). Individuals with spinal cord injuries, traumatic brain injuries, muscular dystrophies, strokes, Rett syndrome, and other neurological conditions commonly use Manual Standing wheelchairs (16). Manual standing wheelchairs enable users to maintain standing positions and provide adequate support for their ankles, knees, and hips (6).

A standing wheelchair is designed to meet the medical and functional needs of individuals with disabilities (17). It also improves functional reach, mobility, circulation, and range of motion, and reduces contracture risk in individuals with lower limb paralysis. Along with many other psychosocial and quality-of-life advantages, standing wheelchairs improve bone health, and vital organ capacity, and reduce muscle tone, pressure ulcers, and skeletal deformities (18). However, existing standing wheelchair designs are costly due to the use of high technology electrical and electronics and which require added maintenance (19). Additionally, it is becoming less reliable due to the electronics involved. Moreover, access to therapy or regular standing programs is severely hampered for people who are economically challenged or live far away.

To address this, we have recently developed a low-cost standing wheelchair from locally available materials in Dhaka, Bangladesh. The wheelchair was tested by individuals with SCI at the Centre for Rehabilitation of the Paralyzed, Savar, Bangladesh. In this report, we have shared the design which has also been published on an open-source repository and summarized the results of the test on returning independence and enhancing the life quality of these individuals with paralysis. Our results suggest that the standing wheelchair significantly enhances individuals’ overall quality of life (QoL).

Our developed low-cost open-source standing wheelchair has the potential to bridge the 10:90 gap (20) and potentially decrease the disparity of assistive devices worldwide (21).

## DESIGN

The design process of the standing wheelchair involved the active participation of designers, rehabilitation engineers and rehabilitation specialists including clinical physiotherapists. ***Figure 1*** shows the complete design of a low-cost standing wheelchair. The entire design and source files are freely shared in an online repository: https://github.com/OpenMedTech-Lab/OpenXwheel

**Figure 1:**
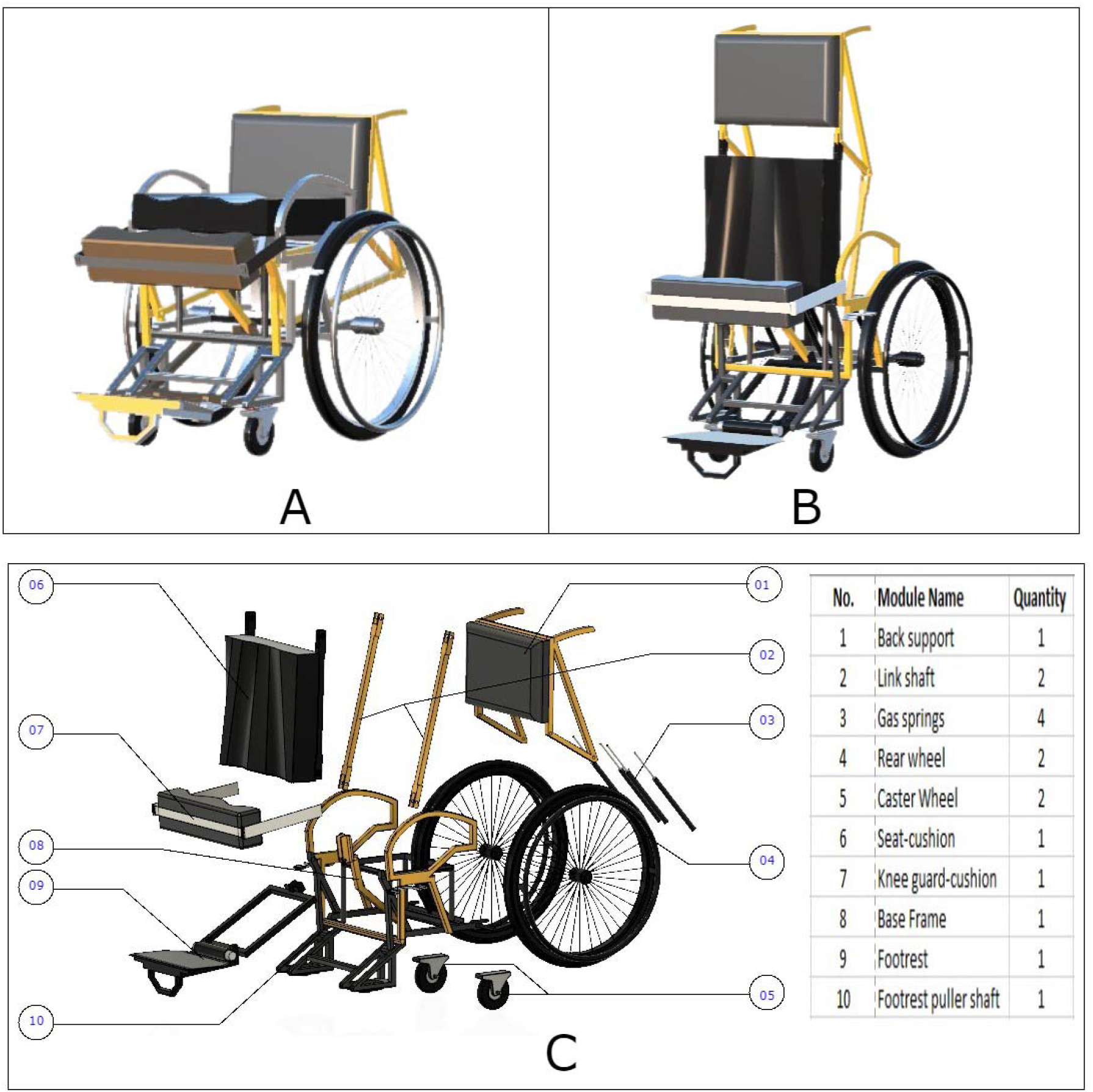
Open-source design of a low-cost standing wheelchair: **A)** sitting **B)** standing **C)** exploded view of different components of the wheelchair.

The standing wheelchair was prototyped following the existing wheelchair model of CRP and in accordance to the guideline of the world health organization (WHO) (19). The standing wheelchair comprises a total of 9 components: seat, base, back-support, armrest, footrest, rear wheel, castor wheel, push rim, brakes, chest support, and knee block. The wheelchair’s main frame holds all parts together which makes it economical to repair.

The back of a standing wheelchair is composed of a cushioned board, and the base is a horizontal board with a contour cushion. The contour cushion was employed to prevent pressure ulcers (22). A wide chest belt was utilized to prevent any potential falls. To provide knee stability, knee blocks were utilized which also assist in standing upright. An additional bar was added to the footplate to prevent forward falling. Four brakes were used in a standing wheelchair, including two hand brakes for securing the wheelchair and two brakes for ensuring safe sitting and standing positions.

Link shafts are essential components in ensuring the chair’s seat and back support remain erect during its standing position. Two rear and caster wheels (Dhaka Cycle Store, Bangladesh) were utilized for smooth movement in both outdoor and indoor environments.

We utilized Nitrogen oxide gas spring mechanisms (Automobile Soring House, Bangladesh) in our standing wheelchair to minimize user effort and assist them in converting from a sitting to a standing position. Autodesk Fusion 360 was utilized during the design development stage to analyze stresses in the solid model of the standing wheelchair design. The gas spring applied to an empty standing wheelchair in the upright position generated significant forces at the joints, resulting in maximum forces experienced. Before a trial with patients, we conducted several safety tests with able individuals.

## METHODS

The standing wheelchair had undergone multiple refinements from August 2022 to May 2023, incorporating user feedback. The trial was conducted at the Centre for the Rehabilitation of the Paralysed (CRP) and the ethics committee granted permission for the study (CRP-ERC-R&E-0401-0468). All study participants were recruited from CRP’s inpatient SCI unit.

### Subjects

30 SCI paraplegic inpatient wheelchair users (n = 30; 26 Males, 4 Females) were enrolled to test the wheelchair. Purposive sampling was used to select the study participants. The following inclusion and exclusion criteria were used to recruit the study participants:

### Experimental procedure

Written informed consent was obtained from the study participants. The study was conducted over 9 months at the Centre for the Rehabilitation of the Paralysed (CRP). 30 30-minute interactive training sessions were conducted before the trial to ensure optimal operation. The users received training by having multiple actions carried out in an assisted way.

The first standing wheelchair functions were demonstrated in front of participants. An explanation was given of the advantages and potential risks associated with the use of the standing wheelchair. Activities and durations of the tests were also explained. After the training session, the participants were allowed to operate the standing wheelchair independently. The design and clinical teams conducted observations. The testing sessions typically lasted 30 minutes. A wheelchair usability questionnaire, listed in ***Table 2*** was used to collect the data with the use of a 10-point Likert scale and their feedback was obtained through a qualitative questionnaire.

**Table 1:**
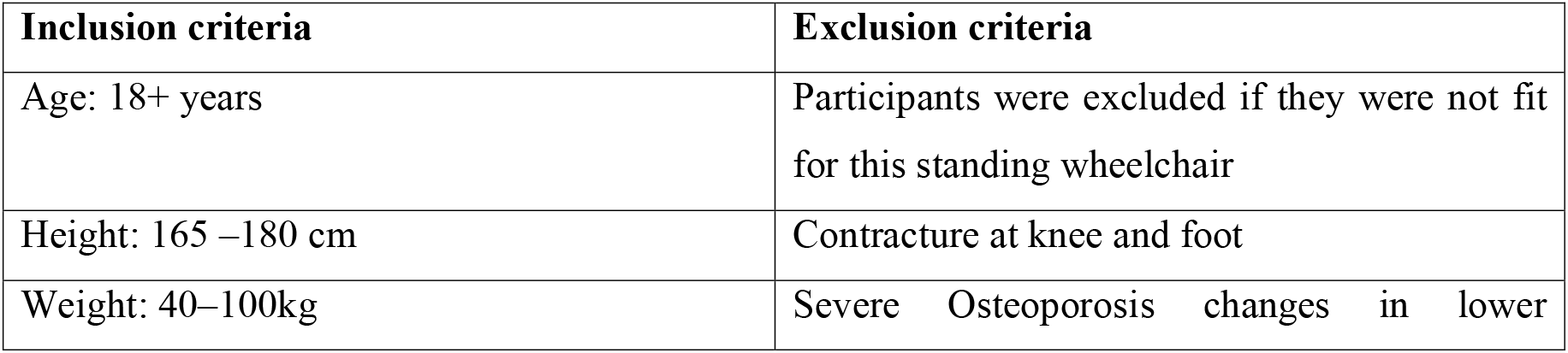
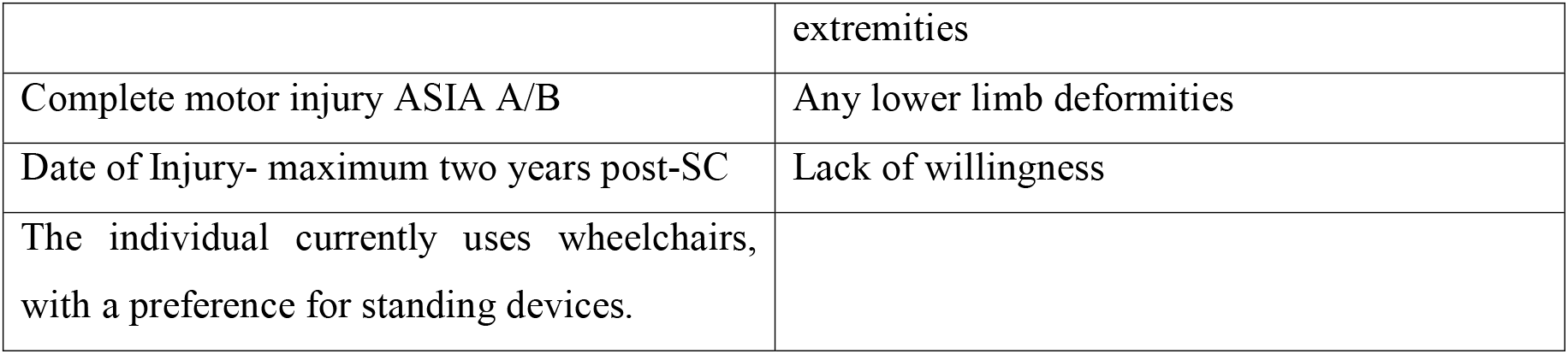
The selection of subjects was based on specific inclusion and exclusion criteria:

**Table 2:** Standing wheelchair usability and acceptability questions:

| **Questionnaire** | **Score** |
| --- | --- |
| 1. How safe do you feel in your new standing wheelchair? | 0-10 (10 is safest) |
| 2. Is it easy or difficult to operate a standing wheelchair? | 0-10 (10 is most difficult) |
| 3. How easy or difficult do you think the new wheelchair is to push and move? | 0-10 (10 is most difficult) |
| 4. How confident would you feel talking to people standing on your own using the standing wheelchair? | 0-10 (10 is most confident) |
| 5. How happy do you feel when you are standing? | 0-10 (10 is most happy) |
| 6. How scared do you feel when you are standing? | 0-10 (10 is most scared) |
| 7. How confident do you think you can do more activity at home if you could stand using the standing wheelchair? | 0-10 (10 is most confident) |
| 8. How would you rate your satisfaction with the standing wheelchair? | 0-10 (10 is highest satisfaction) |
| 9. Is the wheelchair stable and comfortable while sitting and rolling? | Yes/No |
| 10. Is the standing upright and stable while standing using the wheelchair? | Yes/No |
| 11. Would you prefer to use this standing wheelchair instead of your conventional wheelchair? | Yes/No |

**Table 3:** Standing wheelchair fitting and safety Checklist (adopted from the WHO guideline)

|  |  |
| --- | --- |
| Foot position | ☒ |
| Armrest position | ☒ |
| Knee block placement | ☒ |
| Chest belt placement | ☒ |
| Apply and release wheel lock mechanism during sitting | ☒ |
| Apply and release wheel lock mechanism during standing | ☒ |
| Push up on armrest | ☒ |
| Move forward | ☒ |
| Move backward | ☒ |
| Turn right | ☒ |
| Turn left | ☒ |
| Wheel up a slope | ☒ |
| Wheel down a slope | ☒ |
| Sitting position to standing position | ☒ |
| Standing position to sitting position | ☒ |
| Standing mechanism practice | ☒ |
| Try to stretch your hands towards the front, left and right while standing | ☒ |

The standing wheelchair was designed to accommodate users of various sizes and body weights, up to 100 kg. The standing wheelchair was adjusted for each user prior to training or test activities to ensure a proper fit. To achieve a upright standing position, for each study participant, we ensured knee stability with a knee block, use a chest belt for erect posture and unlock the sitting brake.

Once every check was confirmed, as listed in ***Tabel 3***, users push both armrests by themselves to initiate a standing position, which then converts the force into a gas spring to enable them to stand. User then activated a second brake to guarantee safe standing after. For entire process, please see the ***Supplementary Video 1***.

Once trained, no assistance was required for any study participant to stand using our standing wheelchair. All our participants could maintain upright sitting and standing posture and they also preferred it over their conventional wheelchair or a standing frame.

## RESULTS

### The standing wheelchair was found feasible for a wide spectrum of people with SCI

Our trial suggests that the manual standing wheelchair is feasible in a wide spectrum of males and females (26:4) with a wide range of individuals aged 18-49 and mean age of 30 as listed in ***Table 4***. The individuals’ height range was 152.4 to 180.34 inches, and their weight range was 44kg to 75 kg. Most patients were ASIA-A (both motor and sensory complete injury) (18) and 4 being ASIA-B (motor complete and sensory incomplete injury) with neurological levels between T1 to L3. This trial involved participants 1-5 years post-injury (***Figure 2***). The individuals used the wheelchair for an average of 5 to 8 hours per day.

**Table 4:** Demographic characteristics of the study participants:

| Demographic Variables-1 | Mean | Standard- Deviation |
| --- | --- | --- |
| Age (year) | 30.8 | 8.5 |
| Height (inch) | 165.02 | 6.8 |
| Weight (kg) | 57.3 | 8.3 |
| Daily sleep (hours)<sup>‡</sup> | 7.7 | 1.2 |
| **Demographic Variables-2** | **N** | **%** |
| Male | 26 | 86.7 |
| Female | 4 | 13.3 |
| **Clinical Variables** | **N** | **%** |
| ASIA-A | 26 | 86.7 |
| ASIA-B | 4 | 13.3 |
| **Neurological level** |  |  |
| T1-T4 | 3 | 10 |
| T5-T8 | 8 | 26.7 |
| T9-T12 | 16 | 53.3 |
| L1-L3 | 3 | 10 |

**Figure 2:**
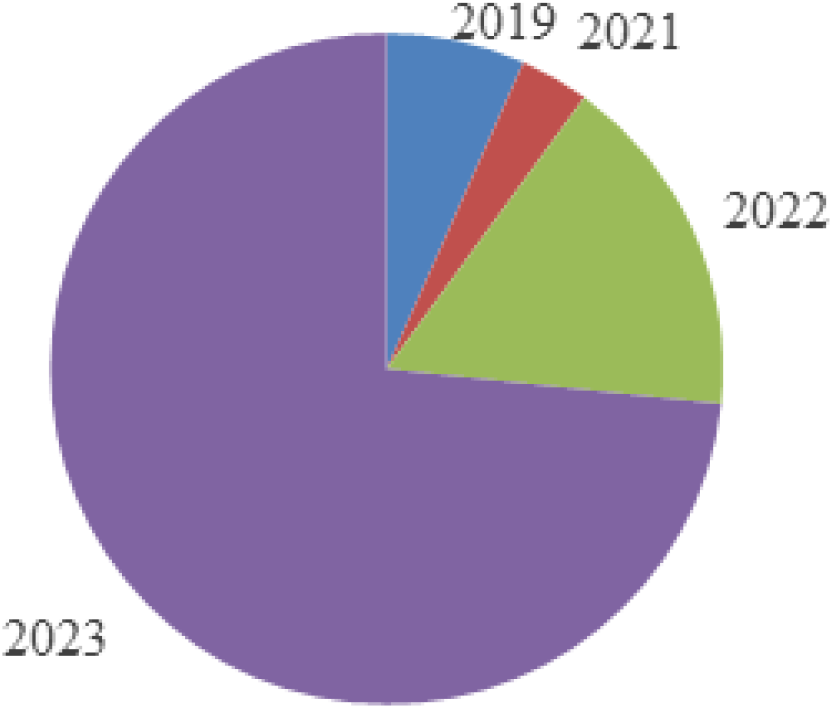
Distribution of participants by year of injury.

### The standing wheelchair was easy to operate for the regular wheelchair users

Being able to talk to people in a standing position using the standing wheelchair made them feel more confident (7.7 / 10). Furthermore, the users also anticipated that the standing wheelchair would enhance their involvement in household activities (7.3 /10). It also improved their confidence level as shown by their responses (***Figure 3***).

**Figure 3:**
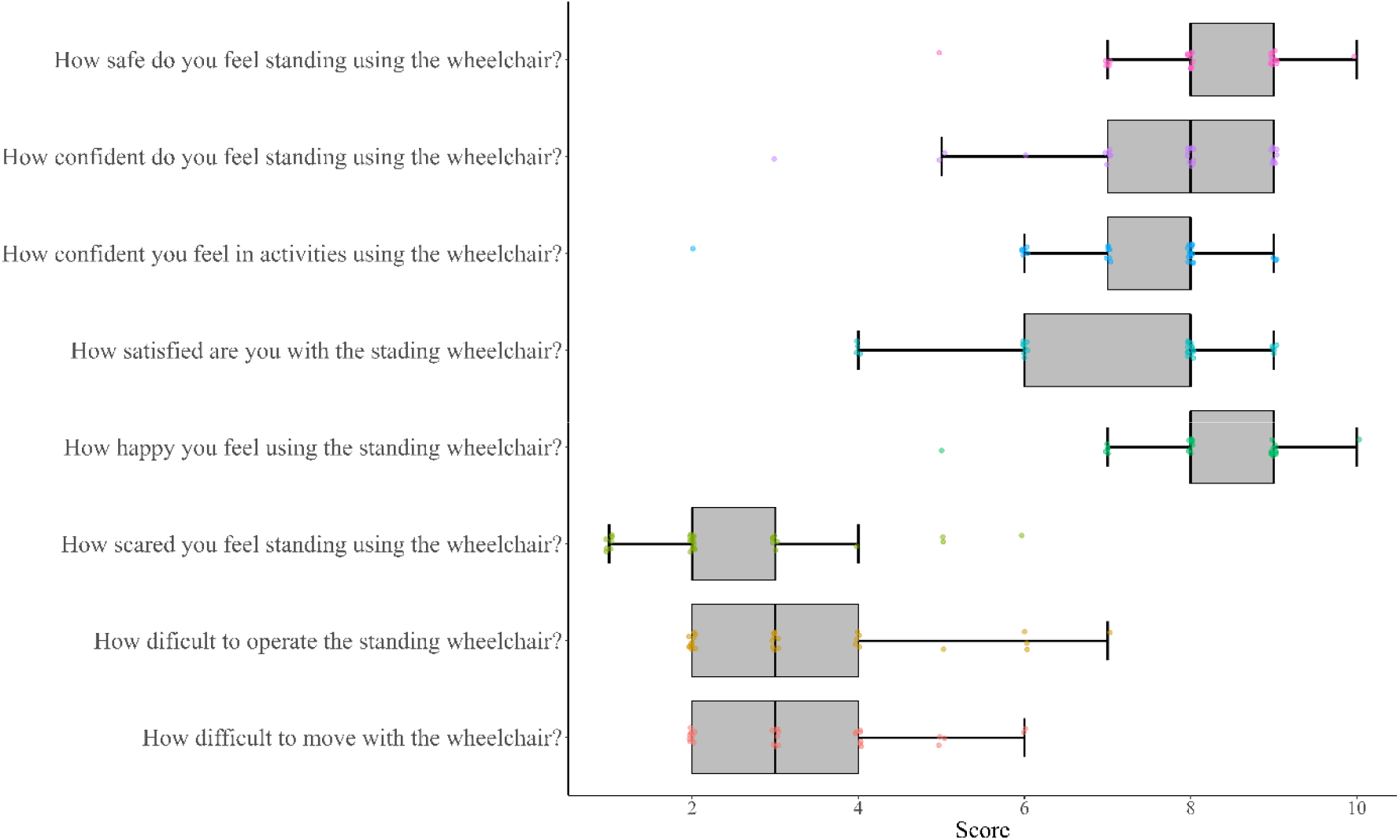
Average responses to the questioners of using the manual standing wheelchair.

### The conventional wheelchair users felt confident and safe using the standing wheelchair

Our questionnaire data suggest that the study participants felt safe (8.4/10) in our standing wheelchair and all subjects showed a lower level of nervousness (2.4 / 10) which significantly indicates that our standing wheelchair is safe for the patients. Very little efforts were needed to operate and propel (3.3 / 10) the wheelchair, and these criteria make sure that our manual standing wheelchair is highly acceptable among regular wheelchair users.

### The wheelchair users felt satisfied and happy using the standing wheelchair

Our participants felt very happy (8.2 / 10) and the overall user satisfaction of the standing wheelchair was high (7.1 / 10). In summary, our developed standing wheelchair offers standing functionality with simplicity of operation. Our prototype also appears feasible and affordable (US$166) in the context of the developing world.

## DISCUSSION

The paper describes the development of a standing wheelchair that allows a paraplegic to maintain a standing position. The prototype standing wheelchair enables users to effortlessly move from sitting to standing and back, utilizing their energy and body weight without relying on any electrical or electronic components. We have collected the user’s feedback in response to our prototype. The objective of this study was to improve the feasibility, usability and acceptability of a low-cost manual standing wheelchair. Since it is completely manual, reliable, user-friendly, dependable, and economical further improvements are needed for complete development.

The manual standing wheelchair underwent several design reviews throughout its development. The design process required multiple analyses to ensure user needs, safety, strength, and manufacturability were met. During the design process development stage, previous work was observed, consumer and clinician requirements were identified, and a pilot model was developed. The goal is to create a wheelchair with a hand-powered standing mechanism that is affordable, easy to repair, and functional, enabling users to engage in economic activities. All the design files and technical details can be downloaded from the OpenMedTech-Lab website (https://openmedtech-lab.github.io/). This open-source approach is a significant step toward making this high demand assistive device more affordable and accessible, particularly in resource-limited settings.

Our developed manual standing wheelchair was not adjustable, but customization is a crucial aspect of an assistive device. Further development is required according to ISO and WHO’s guidelines, which will be adjustable. The trial was conducted exclusively for participants with SCI in a hospital setting, and future trials should be conducted with other neurological conditions in a community setting. The standing wheelchair’s weight exceeds that of a conventional wheelchair, prompting a plan to decrease its weight in the next prototype. Our future project aims to motorize standing wheelchairs for tetraplegic patients, enabling them to stand with minimal effort.

The standing wheelchair would change the lives of many individuals around the world by improving their physical health, mental health as well overall QoL and will help to meet the WHO’s 2030 target. This low-cost feature increases SWC accessibility to a larger population of underserved people with similar disabilities. However, it would be beneficial to conduct further testing in non-hospital settings. Future studies are required to evaluate the potential benefits of a standing wheelchair to prevent secondary complications particularly pressure ulcers after SCI.

## Data Availability

All data produced in the present study are available upon reasonable request to the authors

## ACKNOWLEDGEMENTS

We sincerely thank all the study participants for their time, valuable feedback, and contribution to the evaluation of the manual standing wheelchair.

